# Who leads Marburg virus research? Gender, geography and funding in authorship, 2008 to early 2023: a bibliometric baseline before the 2023–2025 outbreaks

**DOI:** 10.64898/2026.11.04.26367239

**Authors:** Maxime Descartes Mbogning Fonkou

**Author notes:** **Correspondence:** Maxime Descartes Mbogning Fonkou. **Code, data and interactive explorer:** https://github.com/descartesmbogning/marburg-equity https://cognosx.ca/dashboards/marburg-equity.html.

## Abstract

**Introduction:** Between 2023 and 2025, Marburg virus disease reached Equatorial Guinea, Tanzania, Rwanda and Ethiopia for the first time. WHO’s 2025 Marburg research priorities make leadership of studies by local scientists a continuing goal. This study describes who led Marburg virus research before these outbreaks.

**Methods:** I analysed 684 English-language Web of Science papers on Marburg virus published from 2008 to February 2023 (5,700 authorships). Gender was inferred from first names with Gender-API, papers about African countries were identified from their text, funders were harmonised by country and type, and themes were mapped to the WHO priorities. I used Wilson 95% confidence intervals (CIs) and logistic and negative binomial regression.

**Results:** Women held 35.1% (95% CI 33.9–36.4) of authorships, 41.6% (37.9–45.4) of first-author and 19.6% (16.7–22.9) of last-author positions. Women last authors rose from 15.1% (2008–2012) to 23.2% (2018–2023). Only 14.6% of papers had an author based in Africa. Of 102 papers about an African country, 25.5% (18.0–34.7) had a first author and 23.5% (16.4–32.6) a last author from that country, below benchmarks for infectious-disease research in Africa. Women based in Africa were last author on 2 papers. Of funded papers, 69.0% acknowledged a US funder (24.0% a US defence agency) and 3.2% an African funder. WHO priorities on clinical care, outbreak trials, social science and capacity strengthening each appeared in fewer than 8% of papers. An apparent citation penalty for funded papers disappeared after adjustment (rate ratio 1.07, 0.87–1.33).

**Conclusions:** Before the recent outbreaks, Marburg virus research was led mostly by men in senior positions and by researchers outside the affected countries, and was funded mainly by US agencies. This baseline can track whether recent African-led responses change who leads the field.

**Key messages:** *What is already known on this topic:* Marburg virus research has been mapped by output, country and keyword, and the United States dominates it. In infectious-disease and COVID-19 research, women and researchers from African countries are under-represented in senior author positions.

*What this study adds:* To my knowledge, this is the first study of who leads Marburg virus research. Before the 2023–2025 outbreaks, women held one in five last-author positions, only one in four papers about an African country had a first author from that country, and 69% of funded papers acknowledged a US funder. WHO priorities on clinical care, outbreak trials, social science and capacity were each covered by fewer than 8% of papers.

*How this study might affect research, practice or policy:* The findings give a baseline to track whether recent African-led outbreak responses change who leads the field. Funders can direct support to institutions in affected countries and to neglected WHO priorities, and journals can expect local leadership of research done in those countries.

## Introduction

Marburg virus disease is a severe filovirus infection with no licensed vaccine or treatment. Its natural reservoir is the Egyptian fruit bat, and human outbreaks have been rare. They are now becoming more frequent and are reaching new places. Equatorial Guinea and Tanzania reported their first outbreaks in 2023, Rwanda in 2024 and Ethiopia in 2025, and Tanzania had a second outbreak in 2025 [1–5]. Each of these countries had produced little Marburg research before.

The response to these outbreaks also differed from the past. In Rwanda, the national health authorities started a phase 2 vaccine trial ten days after the outbreak was declared and published the main clinical report of the outbreak [3,6]. In Ethiopia, a national research institute provided scientific leadership for the vaccine trial [7]. Filoviruses remain on the WHO list of priority pathogens [8], and in January 2025 the WHO-coordinated Filovirus Collaborative Open Research Consortium published ten Marburg research priorities. These priorities call for equitable participation of researchers from places where the virus circulates and state that leadership of studies by local scientists should be a continuous goal [9].

Several bibliometric studies of Marburg research appeared between 2023 and 2026 [10–14]. They map publication output, journals, countries, keywords and collaboration networks, and they agree that the United States dominates the field. The most recent notes that endemic African countries are under- represented in collaboration networks [14]. None of them asks who leads the work: whether women hold senior author positions, whether researchers from affected countries lead studies about their own countries, or who pays for the research.

These questions have been studied in other fields. Women are under-represented as last authors in biomedical and COVID-19 research [15–17], and the gap is wide in health research from sub-Saharan Africa [18]. In infectious-disease research done in Africa, about half of first authors and four in ten last authors are African [19], and local researchers are often placed in middle author positions or left out altogether [20,21]. During the 2013–2016 Ebola epidemic, only 13% of authors writing about the epidemic were based in Africa [22]. No comparable analysis exists for Marburg virus.

This study provides a baseline for Marburg virus research from 2008 to early 2023, just before the recent outbreaks. It asks four questions: (1) how often women are first and last authors, and whether this has changed; (2) how often researchers from the African countries studied lead research about those countries; (3) who funds the research, and whether the funder’s country is associated with who leads it; and (4) how well research themes matched the WHO 2025 Marburg priorities. It also examines a common pitfall in studies of funding acknowledgements: comparing funded and unfunded papers across years in which funding data were not yet indexed.

## Methods

### Study design and data source

This is a cross-sectional bibliometric study. I searched the Web of Science Core Collection (WoS) on 23 February 2023 with the query Marburg* AND (virus* OR disease* OR hemorrhagic OR haemorrhagic OR viral* OR MARV OR fever) and kept English-language articles, reviews and letters. The search returned 932 records published between 1968 and 2023 (Supplementary Figure S1).

The analysis was restricted to the 684 papers published from 2008 onward, for two reasons. First, WoS has indexed funding acknowledgements systematically only since 2008 [23,24]: in these data, 1.7% of papers published before 2005 had a funding record, compared with 80–86% from 2011 (Supplementary Figure S2). Second, author first names, which gender inference requires, are rarely available in WoS before 2007. Five early-access records without a publication year were assigned their early-access year (2022). The year 2023 is partial and contains 3 papers; it is grouped with the period 2018–2023.

### Authors, gender and author position

Each author was linked to their affiliations through the bracketed author lists in the WoS address field. Gender was inferred from first names with Gender-API, which had the lowest error rate in an independent benchmark of name-to-gender services [25]. Of 5,700 authorships, 96.4% were classified as woman or man; the remainder were left unclassified and excluded from gender denominators. The first author was the first listed author. The last author was the final author on papers with two or more authors and was taken as a proxy for the senior or supervising author. Shares are reported per authorship and by position.

### Geography and local leadership

Each author’s country was taken from the last segment of their address and grouped by World Bank region and FY2023 income group [26]. A paper was classed as being about an African country when that country was named in its title, abstract or author keywords. Virus and strain names that contain place names (for example Sudan virus, Zaire ebolavirus or MARV-Angola) were removed before matching, and ambiguous names required a place context (for example “in Angola” or the province name “Uíge”). A first or last author was counted as local when affiliated in a country the paper was about, and as African when affiliated in any African country. Results were compared with a systematic review of 1,182 papers on infectious-disease research done in Africa [19].

### Funders

Funder names from the WoS “Funding Name Preferred” field were grouped with 54 ordered pattern rules into funder groups, such as the US National Institutes of Health (NIH), the US Department of Defense or the German Research Foundation. Each group was assigned a country and a type: government, government (defence), philanthropy, multilateral, industry or academic. The rules classified 86% of paper–funder links; the remaining 14% were mostly one-off names and were kept as unclassified. The rules are published with the code.

### Research themes and WHO priorities

Research topics were identified with latent Dirichlet allocation fitted to titles, abstracts and author keywords. Missing abstracts were left empty rather than filled with placeholder text. The number of topics (k = 10) was chosen as the value with the highest c_v coherence between 6 and 14 [27], and topics were labelled from their top words. Separately, each paper was tagged against the ten WHO 2025 Marburg research priorities [9] and two themes that are not WHO priorities (basic molecular virology, and biosecurity and biodefence), using a published keyword codebook. A paper could match several themes.

### Citations and open access

Citation counts are WoS Core Collection counts at the date of export. To compare papers of different ages, year-normalised citations were calculated as each paper’s citations divided by the mean citations of papers published in the same year [28]. Open-access status was taken from WoS.

### Statistical analysis

Proportions are reported with Wilson 95% CIs and compared with chi-square tests, or Fisher’s exact test when expected cell counts were small. Logistic regression modelled the odds that the first or the last author was a woman, adjusted for publication year, the region of that author, team size (log number of authors), document type and topic. Negative binomial regression modelled citation counts, with publication-year fixed effects and adjustment for funding, open access, team size, international collaboration, document type and topic. Logistic regression modelled open access by funding status with the same covariates. Subgroup comparisons by topic, region and theme are descriptive; they were not corrected for multiple testing and should be read as exploratory. Analyses used Python (pandas, statsmodels, scikit-learn and gensim). All code, reference tables and a de-identified paper-level dataset are public (see Data and code availability).

### Sensitivity analyses

The gender analysis was repeated in four ways: keeping only Gender-API assignments with an accuracy of at least 80% (89.8% of classified authorships); excluding authors based in East Asia and the Pacific, where name-based inference is least accurate [25]; and counting all unclassified authors first as women and then as men. The local-leadership analysis was repeated for the stricter subset of papers that name the study country in the title.

## Results

### Characteristics of the corpus

The 684 papers had 5,700 authorships from 81 countries. Papers had a mean of 8.3 authors, 1.7 countries and 3.4 unique organisations, and 36.3% involved authors from more than one country. A US-based author appeared on 70.5% of papers. Reviews made up 18.0% of papers. Overall, 81.1% of papers had a funding acknowledgement and 81.3% were open access.

### Gender

Women held a third of all authorships but a fifth of last-author positions (Figure 1; Supplementary Table S1). They held 35.1% (95% CI 33.9–36.4) of the 5,497 classified authorships, 41.6% (37.9–45.4) of 658 first-author positions and 19.6% (16.7–22.9) of 642 last-author positions.

**Figure 1.**
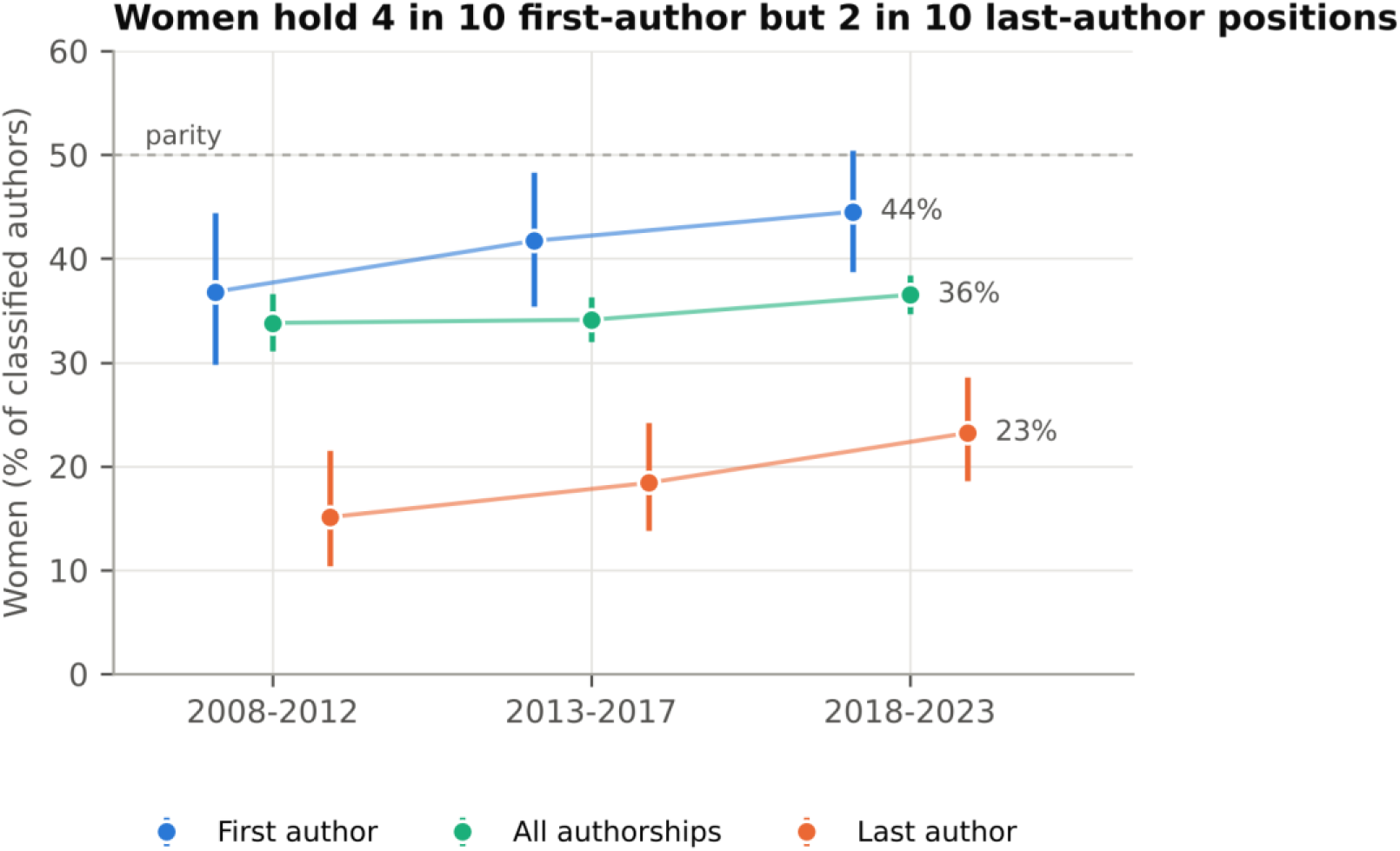
Women by author position and period, 2008 to early 2023. Points show the percentage of women among authors with a classified gender; bars are Wilson 95% confidence intervals. n = 5,497 classified authorships, 658 first authors and 642 last authors.

The share of women last authors rose from 15.1% in 2008–2012 to 18.4% in 2013–2017 and 23.2% in 2018–2023. After adjustment, the odds that the last author was a woman rose by about 6% per year (odds ratio [OR] 1.06, 95% CI 1.01–1.12). The share of women first authors rose from 36.8% to 44.5% over the same periods, but the adjusted trend was not statistically significant (OR 1.04, 1.00–1.09).

Of 628 multi-author papers with both positions classified, 48.1% had a man as first and last author, 32.2% a woman first and a man last, 10.2% a woman in both positions and 9.6% a man first and a woman last.

When the last author was a woman, the first author was also a woman in 51.6% of papers, compared with 40.1% when the last author was a man (chi-square p = 0.026).

The gender gap differed by region. Women were 36.9% of authors based in North America and 40.5% in Europe and Central Asia, but 28.0% in East Asia and the Pacific and 20.1% in sub-Saharan Africa. Only 2 of 25 last authors based in sub-Saharan Africa were women (8.0%). Women were 12.1% of authors based in lower-middle-income countries and 17.1% in low-income countries. In adjusted models, first authors based in East Asia and the Pacific were less likely to be women than those in North America (OR 0.40, 0.21–0.79), and reviews were more likely than other papers to have a woman as last author (OR 3.24, 1.78–5.90).

The results were robust to the four sensitivity analyses of gender inference (Supplementary Table S5). The share of women last authors ranged from 17.7% when only high-accuracy assignments were kept to 21.8% when all unclassified authors were counted as women.

### Local and African leadership

Researchers based in Africa were rare across the corpus. Only 14.6% of papers had any author based in Africa, 6.4% an African first author and 6.6% an African last author.

Of the 102 papers about an African country, about one in four was led from the country studied (Table 1; Figure 2). Half had at least one author from the country studied, but only 25.5% had a first author and 23.5% a last author from that country. African authorship of these papers was well below the benchmarks for infectious-disease research done in Africa: 29.4% had an African first author (benchmark 49.8%) and 28.4% an African last author (benchmark 41.3%).

**Figure 2.**
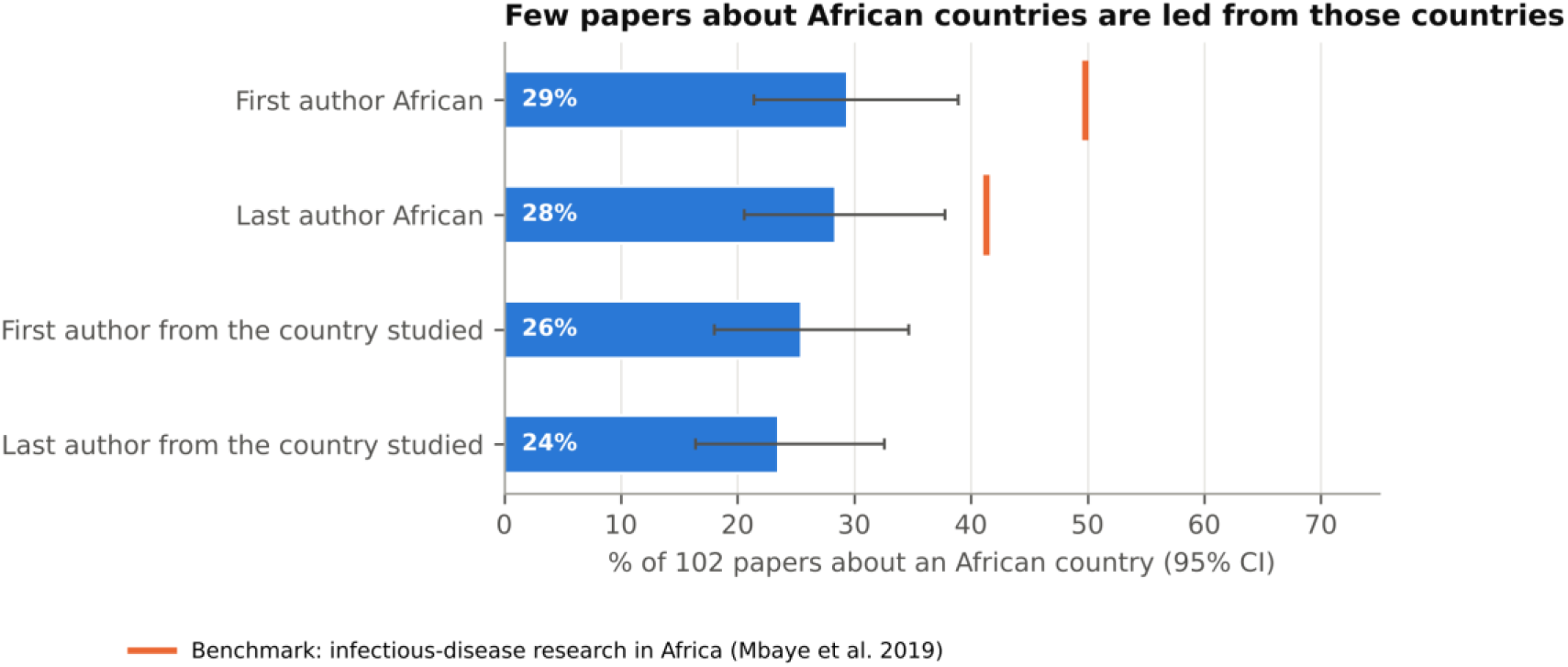
Leadership of the 102 papers about an African country. Bars show the share of papers whose first or last author is based in the country studied or in any African country; whiskers are Wilson 95% confidence intervals. Orange marks are the benchmarks for infectious-disease research done in Africa [19].

**Table 1.** Local and African leadership of 102 papers about an African country, 2008 to early 2023.

| Measure | % (95% CI) | Benchmark, % [19] |
|---|---|---|
| *Author from the country studied* |  |  |
| Any author | 49.0 (39.5–58.6) | – |
| First author | 25.5 (18.0–34.7) | – |
| Last author | 23.5 (16.4–32.6) | – |
| *Author from any African country* |  |  |
| Any author | 61.8 (52.1–70.6) | 93.2 |
| First author | 29.4 (21.4–38.9) | 49.8 |
| Last author | 28.4 (20.6–37.8) | 41.3 |

*The benchmark review reported African authorship only, so no benchmark exists for authors from the country studied. CI: Wilson 95% confidence interval.*

In the stricter subset of 50 papers that name the study country in their title, local first and last authorship were higher (46.0% and 42.0%), and African first authorship (50.0%) matched the benchmark. Even in this subset, most papers were not led from the country studied. African first authorship of papers about African countries rose over time, from 13.0% (3 of 23 papers) in 2008–2012 to 30.8% (8 of 26) in 2013–2017 and 35.8% (19 of 53) in 2018–2023. Across all 684 papers, women based in Africa were first author on 10 papers and last author on 2.

### Funders

Of the 555 funded papers, 69.0% acknowledged a US funder. The US NIH appeared on 47.2% of funded papers and the US Department of Defense on 23.1% (24.0% with other US defence agencies). Funders from Germany (7.7%), Japan (6.5%), Canada (5.6%), China (5.6%), the United Kingdom (5.0%) and the European Union (4.1%) followed. African funders appeared on 3.2% of funded papers (18 papers), and industry on 5.6%.

Funders mostly supported last authors in their own region (Figure 3). Of the 383 papers with a US funder, 84% had a last author based in North America and 3% a last author in sub-Saharan Africa. Of the 18 papers with an African funder, half had a last author based in sub-Saharan Africa. Of the 57 papers with an African first or last author, 77.2% were funded: 26.3% by an African funder and 36.8% by a US funder.

**Figure 3.**
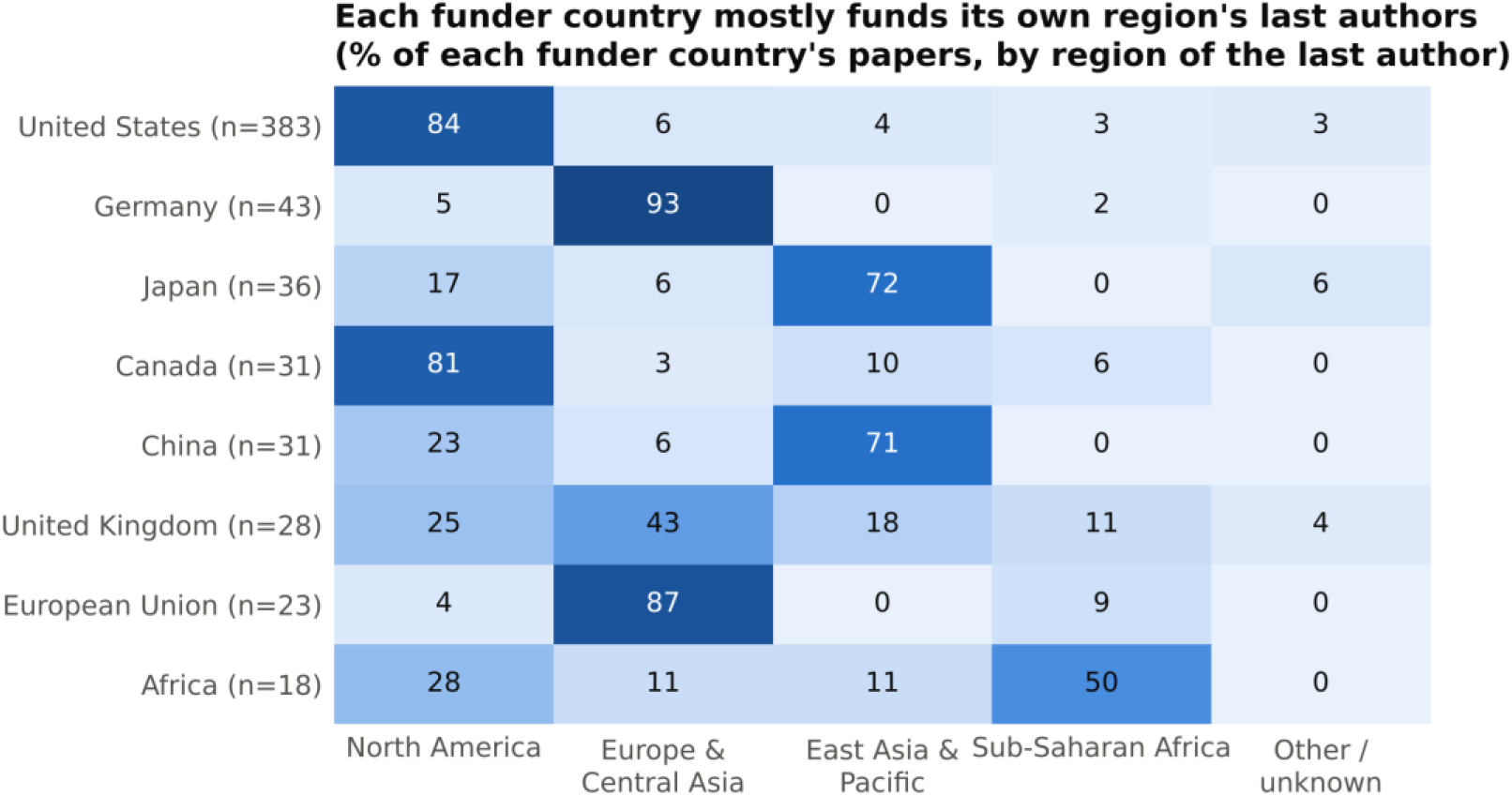
Region of the last author by funder country. Each row sums to 100% of the papers that acknowledge a funder from that country (n in brackets). A paper with funders from several countries appears in each row.

### Research themes and WHO priorities

The ten topics ranged in size from vaccines and animal challenge models (123 papers) to innate immunity and interferon antagonism (23 papers). Authors based in Africa appeared on 32.9% of papers on outbreaks and public health, 31.2% of diagnostics papers and 27.6% of papers on bat reservoirs, but on 1.4% of papers on viral assembly and on none of the papers on innate immunity. Women were first authors on 53.4% of viral assembly papers but on 21.0% of overview papers.

Research concentrated on medical countermeasures (60.1% of papers) and basic molecular virology (59.6%) (Figure 4). Four WHO priorities were each covered by fewer than 8% of papers: supportive clinical care and infection prevention (5.3%), research during outbreaks, including trials (3.9%), social sciences and community engagement (7.2%), and collaboration and capacity strengthening (6.0%).

**Figure 4.**
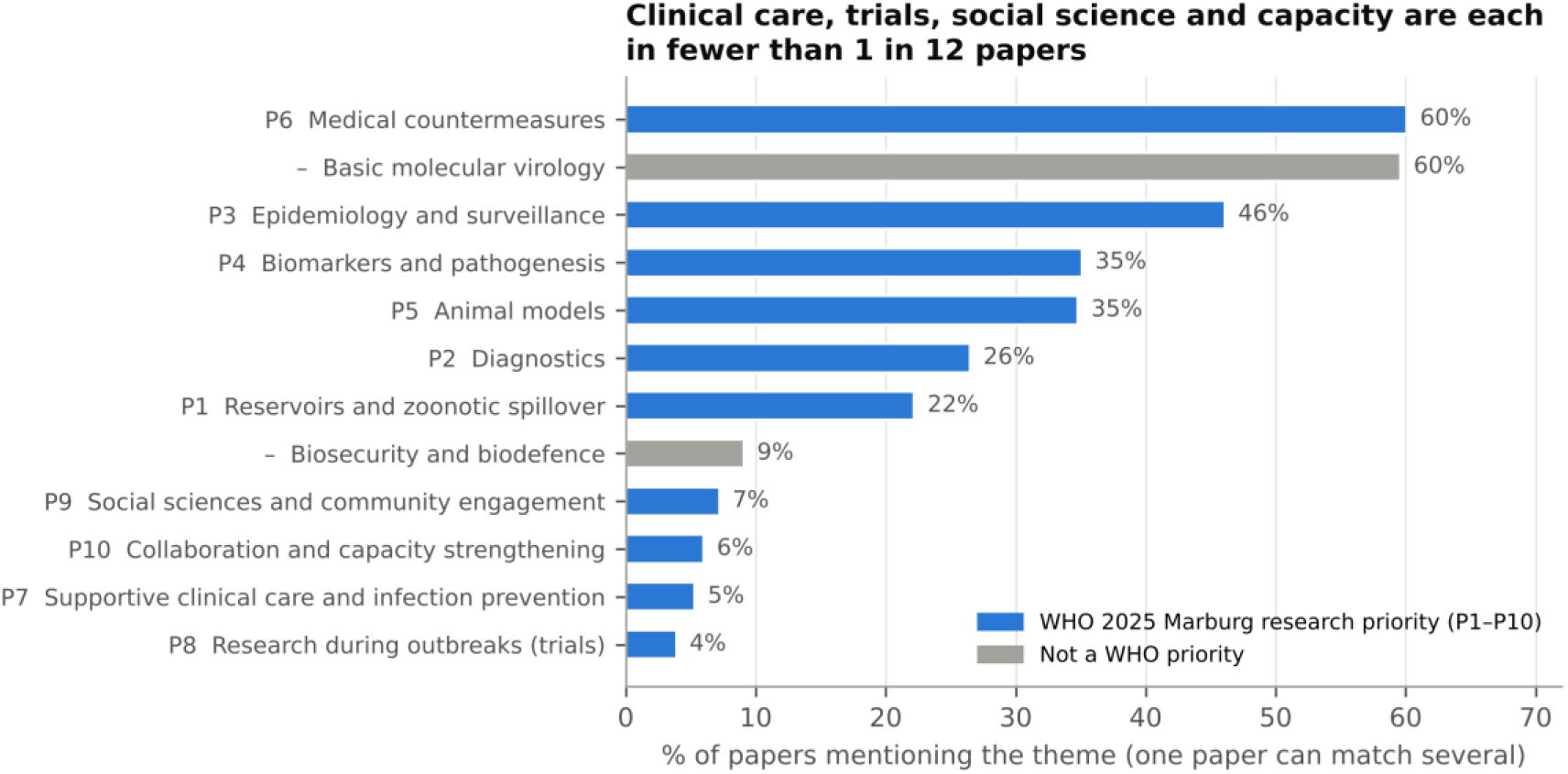
Coverage of the WHO 2025 Marburg research priorities. Share of the 684 papers that match each theme’s keyword codebook. A paper can match several themes. Grey bars are themes that are not WHO priorities.

The thematic profile differed by funder and by leadership. Papers funded by US defence agencies were mostly about medical countermeasures (76.7%) and animal models (48.9%), and rarely about social science (3.0%) or capacity strengthening (0.8%). Papers with an African first or last author more often covered epidemiology and surveillance (70.2%), reservoirs (56.1%), social science (21.1%), capacity strengthening (19.3%) and clinical care (10.5%).

### Funding, citations and open access

Across all 932 records from 1968 onward, papers with a funding acknowledgement had fewer mean citations than papers without one (38.7 vs 55.8; Mann–Whitney p < 0.001), a comparison often reported in studies of research funding. This comparison is confounded by publication age: most “unfunded” papers were published before WoS indexed funding data, so they had more time to collect citations (Supplementary Figure S2). Within 2008–2023, funded papers had higher year-normalised citations than unfunded papers (1.03 vs 0.85).

Regression models within 2008–2023 showed how the apparent effect of funding depends on adjustment (Supplementary Figure S3). Without adjustment, funding was not associated with citations (rate ratio 1.16, 95% CI 0.90–1.48). After adjustment for publication year, funded papers were cited more (1.28, 1.04–1.57). After full adjustment, the association disappeared (1.07, 0.87–1.33). The year-adjusted difference was therefore explained by other characteristics of funded papers, mainly open access: open- access papers were cited more (adjusted rate ratio 1.87), and funded papers were more often open access (adjusted OR 2.34, 1.39–3.96).

Citations also differed by the gender of the first and last authors. Compared with papers with a man in both positions, papers with a woman in both positions received fewer citations (adjusted rate ratio 0.76, 0.59–0.99), and papers with a man first and a woman last received more (1.31, 1.01–1.71). Papers with a woman first and a man last did not differ (0.90, 0.76–1.06).

## Discussion

### Principal findings

In the 15 years of Marburg virus research before the 2023–2025 outbreaks, women held one in five senior author positions, and researchers from the countries studied led about one in four papers about those countries. US government agencies, including defence agencies, funded most of the work. The research centred on countermeasures and basic virology, while the WHO priorities on clinical care, outbreak research, social science and capacity were rarely addressed. Together, these findings describe a field that was largely led from outside the places where the virus circulates, at the moment those places began to face it.

### Gender

The 19.6% share of women last authors is low compared with other fields. It is below the 35% reported for health research from sub-Saharan Africa [18], and it is consistent with the under-representation of women among authors of COVID-19 research [16,17]. The gap between first authorship (42%) and last authorship (20%) follows the pattern seen across science, in which women’s share falls with seniority [15]. The rise in women last authors to 23% in 2018–2023 is encouraging, but it is slow.

Women first authors were more common when the last author was a woman. This is consistent with evidence that women faculty in the life sciences train more women than men faculty do [29]. If this holds in Marburg research, the shortage of senior women may also limit the next generation of women researchers. The citation differences by gender pairing point in different directions and come from small groups, so they should be interpreted with caution. The near-absence of African women in senior positions (2 last-author papers in 15 years) is where the gender gap and the geographic gap meet most clearly.

### Local leadership

African first and last authorship of papers about African countries (29% and 28%) was well below the 50% and 41% found for infectious-disease research done in Africa [19]. By this measure, Marburg virus research was less locally led than infectious-disease research in Africa as a whole. Part of this gap reflects the nature of the field. Laboratory work with live Marburg virus requires biosafety level 4 containment, and of 59 such laboratories worldwide, only 3 are in Africa [30]. Papers based on samples or viral sequences from Africa may therefore be analysed and led elsewhere.

The stricter title-based analysis found higher, but still minority, local leadership, so the main result is not an artefact of loose country tagging. The rise in African first authorship over time, and the locally led responses in Rwanda and Ethiopia [3,6,7], suggest that the field may be changing. This baseline makes it possible to measure that change. Journals and funders can support it: PLOS Medicine, for example, now expects local researchers to be included as authors of research done in their countries [31].

### Funding and the research agenda

The dominance of US government funding agrees with funding-flow data. The G-FINDER survey found that Marburg received about 1.3% of tracked emerging-infectious-disease R&D funding between 2015 and 2022, that the NIH was its largest funder, and that industry provided no funding at all [32]. This study adds two findings. First, a large share of funded papers acknowledged defence agencies, and these papers focused on countermeasures and animal models rather than clinical care, social science or capacity. This pattern fits a biodefence rationale for Marburg research, although the data cannot show whether the funder’s priorities shaped the topics or whether defence agencies funded groups already working on these topics. Second, funders mainly supported last authors in their own region, and only a quarter of African- led papers acknowledged an African funder. Building local leadership will therefore probably need funding that flows directly to institutions in affected countries, and not only to collaborations led from abroad.

### A methodological note on funding acknowledgements

A naive comparison suggests that funded papers are cited less. This result arises from comparing papers across years in which WoS did not yet index funding, so that “unfunded” largely meant “old”. When the analysis was limited to the indexed period and adjusted for publication year and paper characteristics, the effect disappeared. Studies that use WoS funding acknowledgements should restrict their analyses to 2008 onward and normalise citations by year [23,24,28].

### Strengths and limitations

To my knowledge, this is the first study of gender, local leadership and funding in Marburg virus research, and it compares the research directly with the WHO 2025 priorities. The pipeline is reproducible, and the code and de-identified data are public, together with an interactive explorer of the results.

The study has several limitations. WoS misses some regional journals and non-English papers, which may under-count African authors. Gender inference from names is binary, less accurate for some name origins [25] and silent on how people identify; 3.6% of authorships could not be classified, although the sensitivity analyses suggest that this does not change the conclusions. Last authorship is an imperfect proxy for seniority, and shared or alphabetical authorship was not detected. Affiliation is not nationality, so researchers from affected countries who work abroad were counted as non-local. Tagging the study country from text is approximate, even with strain names removed and the stricter title-based check.

WHO priority tags are keyword-based and show broad emphasis rather than precise counts. Funding acknowledgements record what authors declare, not amounts. Finally, the data end in February 2023, before the recent outbreaks, so the results describe a baseline.

### Implications and future research

Three actions follow from these findings. Funders can route support to institutions in affected countries and to the neglected WHO priorities on clinical care, outbreak research, social science and capacity.

Journals can expect local first or last authorship for research done in affected countries. Researchers can repeat this analysis with 2023–2026 data to measure whether the locally led responses in Rwanda and Ethiopia changed who leads the field.

## Conclusion

Before the 2023–2025 outbreaks, the people who led Marburg virus research were mostly men in senior positions, were based outside the countries where the virus circulates, and were funded mainly by US government agencies. The research rarely addressed the clinical, social and capacity priorities that WHO now names. This baseline can be used to measure whether the recent shift towards locally led outbreak research continues.

## Declarations

### Ethics

This study used published bibliographic records only and involved no human participants, so ethics committee approval and consent were not required. Gender was inferred from authors’ first names; it is reported only in aggregate, and the gender of individual authors is not published.

### Data and code availability

Code, reference tables, aggregate results and a de-identified paper-level dataset (WoS accession numbers, DOIs and derived variables) are available at https://github.com/descartesmbogning/marburg-equity (MIT licence for code, CC BY 4.0 for data). An interactive explorer of the aggregate results, with downloadable tables, is available at https://cognosx.ca/dashboards/marburg-equity.html. Raw WoS records cannot be redistributed under the WoS licence, and author-level gender data are not shared because they contain personal names.

### Author contributions

MDMF is the sole author and was responsible for conceptualisation, data curation, formal analysis, methodology, software, visualisation, and writing (original draft, review and editing). He is the guarantor.

## Data Availability

Code, reference tables, aggregate results and a de-identified paper-level dataset (WoS accession numbers, DOIs and derived variables) are available at https://github.com/descartesmbogning/marburg-equity (MIT license for code, CC BY 4.0 for data). An interactive explorer of the aggregate results, with downloadable tables, is available at https://cognosx.ca/dashboards/marburg-equity.html. Raw WoS records cannot be redistributed under the WoS licence, and author-level gender data are not shared because they contain personal names.

https://cognosx.ca/dashboards/marburg-equity.html

## Acknowledgements

None.

## Use of AI tools

An AI assistant (Claude, Anthropic) was used to help review code, check numbers against the analysis outputs and edit the text. The author checked all content and takes full responsibility for it.

---

**Supplementary Figures S1.** (study flow), S2 (funding acknowledgements by period) and S3 (funding and citation models) and Supplementary Tables S1–S10 are in the supplementary file.

