## Supplementary Tables S1-S10, Figures S1-S3, Text S1 for "Who leads Marburg virus research? Gender, geography and funding in authorship, 2008 to early 2023: a bibliometric baseline before the 2023–2025 outbreaks"

*Maxime Descartes Mbogning Fonkou · CognosX, Burnaby, Canada ·*

### Supplementary material

Who leads Marburg virus research? Gender, geography and funding in authorship, 2008–2023. All values come from the public pipeline at github.com/descartesmbogning/marburg-equity (`outputs/tables`).

#### Supplementary Figure S1. Study flow

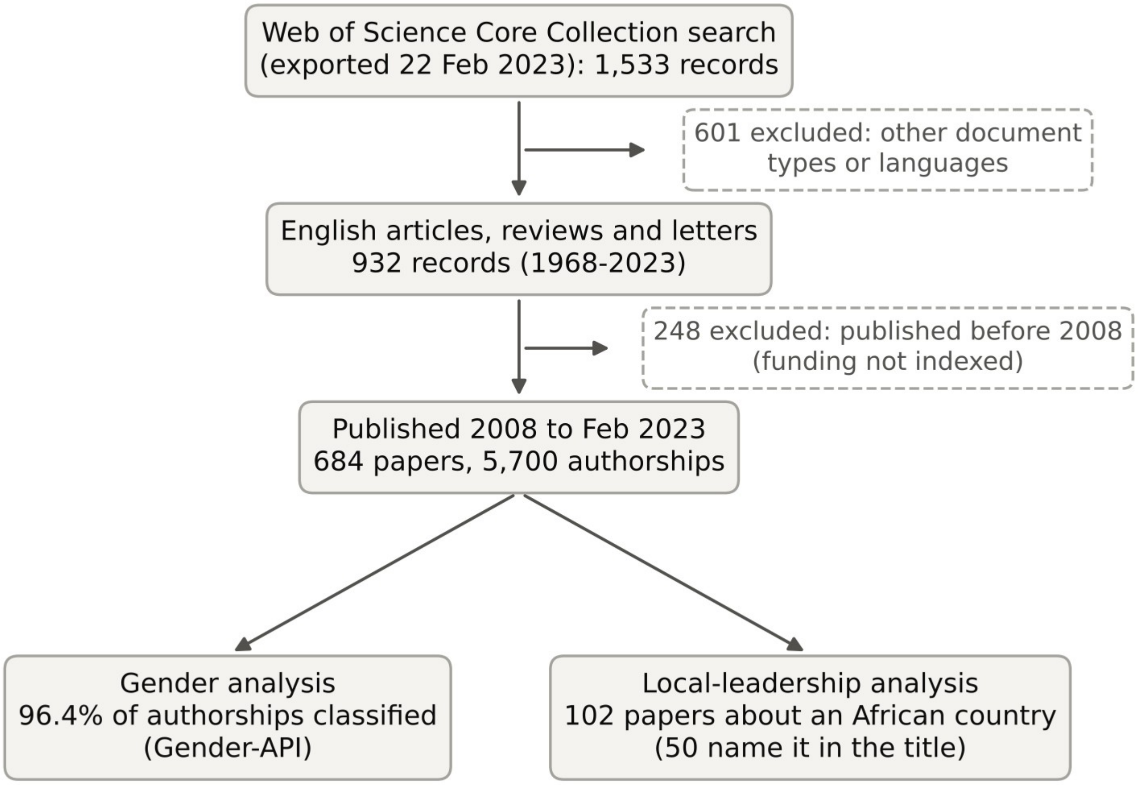

The search returned 1,533 records. Of these, 932 English articles, reviews and letters were kept, and the 684 published from 2008 onward were analysed.

#### Supplementary Table S1. Women by author position, overall and by period

| Position | Women / classified | % women (95% CI) |
|---|---|---|
| All authorships | 1,932 / 5,497 | 35.1 (33.9–36.4) |
| First author | 274 / 658 | 41.6 (37.9–45.4) |
| Last author | 126 / 642 | 19.6 (16.7–22.9) |

| Position | Women / classified | % women (95% CI) |
|---|---|---|
| Middle author | 1,532 / 4,197 | 36.5 (35.1–38.0) |
| Single author | 5 / 24 | 20.8 (9.2–40.5) |

**By period (% women, 95% CI, classified authors)**

| Position | 2008–2012 | 2013–2017 | 2018–2023 |
|---|---|---|---|
| All | 33.8 (31.1–36.6; n=1138) | 34.1 (32.0–36.3; n=1845) | 36.5 (34.7–38.4; n=2514) |
| First | 36.8 (29.8–44.4; n=163) | 41.7 (35.4–48.3; n=223) | 44.5 (38.7–50.4; n=272) |
| Last | 15.1 (10.4–21.5; n=159) | 18.4 (13.8–24.2; n=212) | 23.2 (18.6–28.6; n=271) |

Adjusted odds of a woman last author per year: OR 1.06 (1.01–1.12), p=0.026. First author: OR 1.04 (1.00–1.09), p=0.061 (Supplementary Table S3).

#### Supplementary Table S2. Women by region and income group of the author's affiliation

| World Bank region | Authorships (n) | % women, all | First authors (n) | % women, first | Last authors (n) | % women, last |
|---|---|---|---|---|---|---|
| North America | 3426 | 36.9 | 406 | 43.3 | 396 | 21.0 |
| Europe & Central Asia | 971 | 40.5 | 136 | 47.8 | 131 | 19.8 |
| East Asia & Pacific | 447 | 28.0 | 52 | 26.9 | 62 | 17.7 |
| Sub-Saharan Africa | 417 | 20.1 | 30 | 30.0 | 25 | 8.0 |
| South Asia | 81 | 12.3 | 15 | 20.0 | 12 | 8.3 |
| Middle East & North Africa | 39 | 23.1 | 3 | 0.0 | 5 | 0.0 |
| Latin America & Caribbean | 22 | 40.9 | 4 | 50.0 | 2 | 0.0 |

| World Bank income group (FY2023) | Authorships (n) | % women, all | First authors (n) | % women, first | Last authors (n) | % women, last |
|---|---|---|---|---|---|---|
| High income | 4609 | 36.9 | 559 | 43.6 | 559 | 19.3 |
| Upper middle income | 335 | 36.4 | 47 | 31.9 | 39 | 33.3 |
| Low income | 292 | 17.1 | 16 | 31.2 | 14 | 7.1 |

| World Bank income group (FY2023) | Authorships (n) | % women, all | First authors (n) | % women, first | Last authors (n) | % women, last |
|---|---|---|---|---|---|---|
| Lower middle income | 165 | 12.1 | 24 | 20.8 | 21 | 4.8 |

Groups with fewer than 10 authorships are not shown. Region and income refer to the country of the author's first listed affiliation.

#### Supplementary Table S3. Logistic regression: odds that the first or last author is a woman

**Woman first author**

| Variable | OR (95% CI) | p |
|---|---|---|
| Region: Europe & Central Asia (vs North America) | 1.30 (0.86–1.97) | 0.213 |
| Region: East Asia & Pacific (vs North America) | 0.40 (0.21–0.79) | 0.008 |
| Region: Sub-Saharan Africa (vs North America) | 0.56 (0.24–1.32) | 0.186 |
| Region: Other (vs North America) | 0.64 (0.29–1.43) | 0.278 |
| Topic: Antiviral drugs and entry inhibitors | 1.47 (0.72–3.00) | 0.293 |
| Topic: Bat reservoirs and transmission | 1.69 (0.77–3.70) | 0.189 |
| Topic: Diagnostics and laboratory detection | 1.56 (0.72–3.38) | 0.260 |
| Topic: Filovirus disease overviews | 0.46 (0.20–1.05) | 0.065 |
| Topic: Genome replication and protein structure | 0.86 (0.32–2.33) | 0.762 |
| Topic: Innate immunity and interferon antagonism | 0.92 (0.33–2.59) | 0.874 |
| Topic: Outbreaks and public health risk | 0.79 (0.36–1.74) | 0.554 |
| Topic: Vaccines and animal challenge models | 1.55 (0.79–3.07) | 0.204 |
| Topic: Viral assembly and budding | 2.09 (0.98–4.46) | 0.058 |
| Publication year (per year) | 1.04 (1.00–1.08) | 0.061 |
| Team size (log authors) | 0.96 (0.74–1.23) | 0.732 |
| Review article | 1.03 (0.62–1.72) | 0.902 |

**Woman last author**

| Variable | OR (95% CI) | p |
|---|---|---|
| Region: Europe & Central Asia (vs North America) | 0.89 (0.53–1.50) | 0.663 |
| Region: East Asia & Pacific (vs North America) | 0.71 (0.34–1.46) | 0.347 |
| Region: Sub-Saharan Africa (vs North America) | 0.40 (0.09–1.82) | 0.235 |
| Region: Other (vs North America) | 0.51 (0.16–1.63) | 0.258 |
| Topic: Antiviral drugs and entry inhibitors | 1.46 (0.61–3.45) | 0.393 |
| Topic: Bat reservoirs and transmission | 0.61 (0.21–1.76) | 0.360 |
| Topic: Diagnostics and laboratory detection | 0.88 (0.32–2.37) | 0.793 |
| Topic: Filovirus disease overviews | 0.44 (0.16–1.25) | 0.125 |
| Topic: Genome replication and protein structure | 1.27 (0.40–4.02) | 0.685 |
| Topic: Innate immunity and interferon antagonism | 1.37 (0.41–4.56) | 0.607 |
| Topic: Outbreaks and public health risk | 0.82 (0.31–2.19) | 0.695 |
| Topic: Vaccines and animal challenge models | 0.94 (0.40–2.19) | 0.877 |
| Topic: Viral assembly and budding | 1.05 (0.41–2.75) | 0.914 |
| Publication year (per year) | 1.06 (1.01–1.12) | 0.026 |
| Team size (log authors) | 0.96 (0.67–1.36) | 0.806 |
| Review article | 3.24 (1.78–5.90) | <0.001 |

Reference topic: Antibodies and cross-filovirus immunity.

#### Supplementary Table S4. Gender of first and last authors and citations

| First / last author | Papers | % | Citation rate ratio (95% CI) |
|---|---|---|---|
| Man first / man last | 302 | 48.1 | 1 (reference) |
| Woman first / man last | 202 | 32.2 | 0.90 (0.76–1.06) |
| Woman first / woman last | 64 | 10.2 | 0.76 (0.59–0.98) |
| Man first / woman last | 60 | 9.6 | 1.31 (1.01–1.71) |

The table covers 628 multi-author papers with both positions classified. Rate ratios come from negative binomial regression adjusted for funding, open access, team size, document type and year. Chi-square test of first-author gender by last-author gender: p=0.026.

#### Supplementary Table S5. Sensitivity analyses of gender inference (% women, 95% CI)

| Analysis | All authorships | First author | Last author |
|---|---|---|---|
| Main analysis | 35.1 (33.9–36.4) | 41.6 (37.9–45.4) | 19.6 (16.7–22.9) |
| Gender-API accuracy ≥80% | 34.5 (33.2–35.8) | 41.4 (37.6–45.4) | 17.7 (14.8–21.0) |
| Excluding East Asia & Pacific authors | 35.8 (34.5–37.1) | 42.9 (39.0–46.9) | 19.8 (16.8–23.3) |
| Unclassified counted as women (upper bound) | 37.5 (36.2–38.7) | 43.9 (40.2–47.6) | 21.8 (18.8–25.1) |
| Unclassified counted as men (lower bound) | 33.9 (32.7–35.1) | 40.1 (36.5–43.8) | 19.1 (16.3–22.3) |

89.8% of classified authorships had Gender-API accuracy ≥80%.

#### Supplementary Table S6. Local and African leadership: sensitivity analysis and trend

**Papers naming an African study country in the title (n=50)**

| Measure | Papers | % (95% CI) |
|---|---|---|
| Any local author | 39 | 78.0 (64.8–87.2) |
| First author local | 23 | 46.0 (33.0–59.6) |
| Last author local | 21 | 42.0 (29.4–55.8) |
| First author African | 25 | 50.0 (36.6–63.4) |
| Last author African | 22 | 44.0 (31.2–57.7) |

**Papers about an African country, by period (% and papers)**

| Measure | 2008–2012 | 2013–2017 | 2018–2023 |
|---|---|---|---|
| Any local author | 39.1 (9/23) | 53.8 (14/26) | 50.9 (27/53) |
| First author African | 13.0 (3/23) | 30.8 (8/26) | 35.8 (19/53) |
| Last author African | 13.0 (3/23) | 26.9 (7/26) | 35.8 (19/53) |

Study countries were found in titles, abstracts and author keywords. Virus, strain and disease names that contain place names were removed first (for example Sudan virus, Zaire ebolavirus, Taï Forest virus, Crimean-Congo, MARV-Angola, Egyptian rousette). Sudan, Angola, DR Congo/Zaire and Côte d'Ivoire also required a place context, such as “in Angola”, “Uíge”, “Khartoum” or “Kikwit”.

#### Supplementary Table S7. Funders acknowledged in the 555 funded papers

| Funder group | Country | Type | Papers | % of funded papers |
|---|---|---|---|---|
| US NIH (other institutes) | United States | Government | 175 | 31.5 |
| Other / unclassified | Unknown | Unknown | 144 | 25.9 |
| US NIH (NIAID) | United States | Government | 132 | 23.8 |
| US Department of Defense | United States | Government (defence) | 128 | 23.1 |
| Universities and hospitals | Unknown | Academic | 48 | 8.6 |
| Japanese government | Japan | Government | 36 | 6.5 |
| German Research Foundation (DFG) | Germany | Government | 35 | 6.3 |
| Industry | Unknown | Industry | 31 | 5.6 |
| Chinese government | China | Government | 31 | 5.6 |
| Canadian government | Canada | Government | 31 | 5.6 |
| European Union | European Union | Multilateral | 23 | 4.1 |
| US CDC | United States | Government | 23 | 4.1 |
| German government (other) | Germany | Government | 21 | 3.8 |
| UK government | United Kingdom | Government | 20 | 3.6 |
| African funders | Africa | Government | 18 | 3.2 |
| Japanese foundations | Japan | Philanthropy | 14 | 2.5 |
| Battelle Memorial Institute | United States | Private non-profit | 13 | 2.3 |
| US Department of Energy | United States | Government | 12 | 2.2 |
| Burroughs Wellcome Fund | United States | Philanthropy | 11 | 2.0 |
| Wellcome Trust | United Kingdom | Philanthropy | 11 | 2.0 |

Top 20 groups shown. A paper can acknowledge several funders. 54 pattern rules (`data/reference/funder_rules.csv`) classified 86% of paper–funder links. “Other / unclassified” holds one-off names.

**By funder country**

| Funder country | Papers | % of funded papers |
|---|---|---|
| United States | 383 | 69.0 |
| Unknown (industry, universities, unclassified) | 191 | 34.4 |
| Germany | 43 | 7.7 |
| Japan | 36 | 6.5 |
| Canada | 31 | 5.6 |
| China | 31 | 5.6 |
| United Kingdom | 28 | 5.0 |
| European Union | 23 | 4.1 |
| Africa | 18 | 3.2 |
| International | 12 | 2.2 |
| France | 8 | 1.4 |
| Russia | 6 | 1.1 |

#### Supplementary Table S8. Funding, citations and open access, 2008–2023

##### Negative binomial regression of WoS citations (n=684)

| Model | Rate ratio, funded (95% CI) | p |
|---|---|---|
| M1: no controls | 1.16 (0.90–1.48) | 0.251 |
| M2: + publication year | 1.28 (1.04–1.57) | 0.019 |
| M3: full model | 1.07 (0.87–1.33) | 0.522 |

The full model also includes open access (rate ratio 1.87), team size, international collaboration, document type, year and topic. For comparison, across all 932 records (1968–2023), mean citations were 38.7 for papers with a funding record and 55.8 without (Mann–Whitney $p<0.001$).

##### Logistic regression of open access (n=684)

| Model | OR, funded (95% CI) | p |
|---|---|---|
| No controls | 4.26 (2.78–6.52) | <0.001 |
| Adjusted | 2.34 (1.39–3.96) | 0.001 |

Adjusted for team size, document type, year and topic.

#### Supplementary Table S9. Topic model

##### Model selection (c_v coherence; higher is better)

| Topics (k) | Coherence c_v | Perplexity |
|---|---|---|
| 6 | 0.521 | 1134.1 |
| 7 | 0.561 | 1101.5 |
| 8 | 0.593 | 1094.6 |
| 9 | 0.620 | 1110.0 |
| 10 | 0.642 | 1087.3 |
| 11 | 0.604 | 1076.6 |
| 12 | 0.604 | 1086.6 |
| 13 | 0.623 | 1074.8 |
| 14 | 0.604 | 1095.4 |

**Topics at k = 10**

| # | Topic | Papers | Top words | % women first | % women last | % any African author | % funded | % US defence funded | Closest WHO priority |
|---|---|---|---|---|---|---|---|---|---|
| 0 | Innate immunity and interferon antagonism | 23 | immune, cells, responses, response, infection, ifn, innate, filoviruses, cell, interferon, ebov, activation, type, filovirus, replication | 39.1 | 30.4 | 0.0 | 87.0 | 30.4 | P4 Biomarkers and pathogenesis |
| 1 | Bat reservoirs and transmission | 58 | bats, infection, bat, natural, reservoir, disease, | 48.3 | 13.8 | 27.6 | 84.5 | 27.6 | P1 Reservoirs and zoonotic spillove |

| # | Topic | Papers | Top words | % women first | % women last | % any African author | % funded | % US defence funded | Closest WHO priority |
|---|---|---|---|---|---|---|---|---|---|
| | | | humans, egyptian, host, transmission, aegyptiacus, infected, species, aerosol, rousette | | | | | | r |
| 2 | Outbreaks and public health risk | 79 | health, disease, diseases, outbreaks, outbreak, risk, public, zoonotic, emerging, infectious, fever, control, cases, pathogens, transmission | 30.4 | 19.0 | 32.9 | 55.7 | 3.8 | P3 Epidemiology and surveillance |
| 3 | Antiviral drugs and entry inhibitors | 99 | entry, antiviral, activity, cells, infection, ebov, inhibitors, cell, filoviruses, filovirus, compoun | 42.4 | 23.2 | 6.1 | 90.9 | 20.2 | P6 Medical countermeasures |

| # | Topic | Papers | Top words | % women first | % women last | % any African author | % funded | % US defence funded | Closest WHO priority |
|---|---|---|---|---|---|---|---|---|---|
| | | | ds, receptor, glycoprotein, vitro, compound | | | | | | |
| 4 | Diagnostics and laboratory detection | 64 | fever, assay, hemorrhagic, detection, assays, methods, lassa, samples, analysis, rapid, laboratory, background, diagnostic, specific, developed | 45.3 | 15.6 | 31.2 | 79.7 | 20.3 | P2 Diagnostics |
| 5 | Vaccines and animal challenge models | 123 | vaccine, vaccines, mice, lethal, protection, nonhuman, disease, primates, infection, macaques, challenge, model, efficacy, animals, | 47.2 | 17.9 | 5.7 | 90.2 | 26.8 | P6 Medical countermeasures / P5 Animal models |

| # | Topic | Papers | Top words | % women first | % women last | % any African author | % funded | % US defence funded | Closest WHO priority |
|---|---|---|---|---|---|---|---|---|---|
| | | | filovirus | | | | | | |
| 6 | Viral assembly and budding | 73 | protein, budding, proteins, host, mvp, vlps, particles, domain, membrane, matrix, interactions, assembly, binding, virus-like, egress | 53.4 | 17.8 | 1.4 | 94.5 | 9.6 | Basic virology (no WHO priority) |
| 7 | Antibodies and cross-filovirus immunity | 55 | filovirus, antibodies, ebolavirus, filoviruses, ebov, species, marburgvirus, antibody, sudan, sudv, neutralizing, mabs, ebolaviruses, glycoprotein, human | 34.5 | 18.2 | 25.5 | 87.3 | 30.9 | P6 Medical countermeasures |
| 8 | Genome replication | 29 | rna, genome, structure, replication | 31.0 | 24.1 | 6.9 | 93.1 | 13.8 | Basic virology (no WHO |

| # | Topic | Papers | Top words | % women first | % women last | % any African author | % funded | % US defence funded | Closest WHO priority |
|---|---|---|---|---|---|---|---|---|---|
| | and protein structure | | , filovirus, transcription, region, sequence, sequences, nucleoprotein, complex, gene, proteins, ebov, structural | | | | | | priority) |
| 9 | Filovirus disease overviews | 81 | filovirus, hemorrhagic, fever, disease, development, models, animal, infections, filoviruses, humans, clinical, research, human, vaccines, infection | 21.0 | 13.6 | 9.9 | 56.8 | 16.0 | Mixed (mostly reviews) |

Method: latent Dirichlet allocation (scikit-learn, batch learning, 100 iterations, seed 42) on titles, abstracts and author keywords. The vocabulary kept words that appear in at least 5 papers and in no more than 50% of papers. Each paper is assigned to its dominant topic.

**Supplementary Table S10. WHO 2025 Marburg research priorities: codebook and coverage**

| Code | Theme | Keywords | % all | % 2008–15 | % 2016–23 | % African-led | % US defence funded |
|---|---|---|---|---|---|---|---|
| P1 | Reservoirs and zoonotic spillover | bat(s) · rousettus · reservoir · spillover · zoono* · ecolog* · cave(s) · mine(s) · wildlife · egyptian fruit | 22.2 | 21.0 | 23.2 | 56.1 | 17.3 |
| P2 | Diagnostics | diagnos* · assay(s) · RT-PCR · qPCR · PCR · point-of-care · rapid test · ELISA · lateral flow · detection of | 26.5 | 25.0 | 27.6 | 36.8 | 25.6 |
| P3 | Epidemiology and surveillance | epidemiolog* · surveillance · outbreak · transmission · case fatality · contact tracing · seroprevalence · risk factor | 46.1 | 35.3 | 54.4 | 70.2 | 36.8 |
| P4 | Biomarkers and pathogenesis | pathogenesis · pathogenic · biomarker · cytokine · interferon · innate · host response · immune response · pathology · lesion(s) · coagulopath* · immunopatholog* | 35.1 | 40.0 | 31.2 | 15.8 | 47.4 |
| P5 | Animal models | (non)human primate · NHP(s) · macaque · cynomolgus · rhesus · guinea pig(s) · mice · mouse · hamster · ferret · animal model | 34.8 | 35.7 | 34.1 | 14.0 | 48.9 |
| P6 | Medical countermeasures | vaccin* · monoclonal · mAb(s) · antiviral · inhibitor · drug(s) · remdesivir · favipiravir · therapeutic · treatment · post-exposure · prophyla* · countermeasure | 60.1 | 57.0 | 62.5 | 26.3 | 76.7 |
| P7 | Supportive clinical care and infection | clinical management · supportive care · patient care · intensive care · clinical features · clinical | 5.3 | 5.0 | 5.5 | 10.5 | 1.5 |

| Code | Theme | Keywords | % all | % 2008–15 | % 2016–23 | % African-led | % US defence funded |
|---|---|---|---|---|---|---|---|
| | prevention | presentation · infection prevention · health(care) worker(s) · personal protective · nosocomial · clinical course · case management | | | | | |
| P8 | Research during outbreaks (trials) | clinical trial · ring vaccination · phase 1–3 (I–III) · randomised/randomized · expanded access · trial protocol · compassionate use | 3.9 | 3.7 | 4.2 | 3.5 | 5.3 |
| P9 | Social sciences and community engagement | communit* · social · stigma · anthropolog* · behavio* · perception · awareness · knowledge, attitude · risk communication · trust · qualitative | 7.2 | 7.7 | 6.8 | 21.1 | 3.0 |
| P10 | Collaboration and capacity strengthening | capacity building · capacity strengthening · collaborat* · network(s) of · training program · laboratory capacity · preparedness · one health | 6.0 | 5.3 | 6.5 | 19.3 | 0.8 |
| B1 | Basic molecular virology (not a WHO priority) | VP24, VP30, VP35, VP40 · nucleoprotein · glycoprotein · GP · replication · transcription · budding · genome · crystal structure · polymerase · viral protein · entry · receptor · NPC1 | 59.6 | 61.0 | 58.6 | 43.9 | 64.7 |
| B2 | Biosecurity and biodefence (not a WHO priority) | biosecurity · biodefen* · bioterror* · biological weapon · select agent · biosafety · BSL-4 · category A | 9.1 | 12.3 | 6.5 | 3.5 | 11.3 |

P1–P10 follow the WHO Filovirus CORC top-10 Marburg priorities (January 2025). B1–B2 are not WHO priorities. Matching is case-insensitive on titles, abstracts and author keywords, and a paper can match several themes. "African-led" means the 57 papers with an African first or last author. The exact regular expressions are in `data/reference/who_priorities_codebook.csv`.

##### Supplementary Figure S2. Funding acknowledgements by publication period

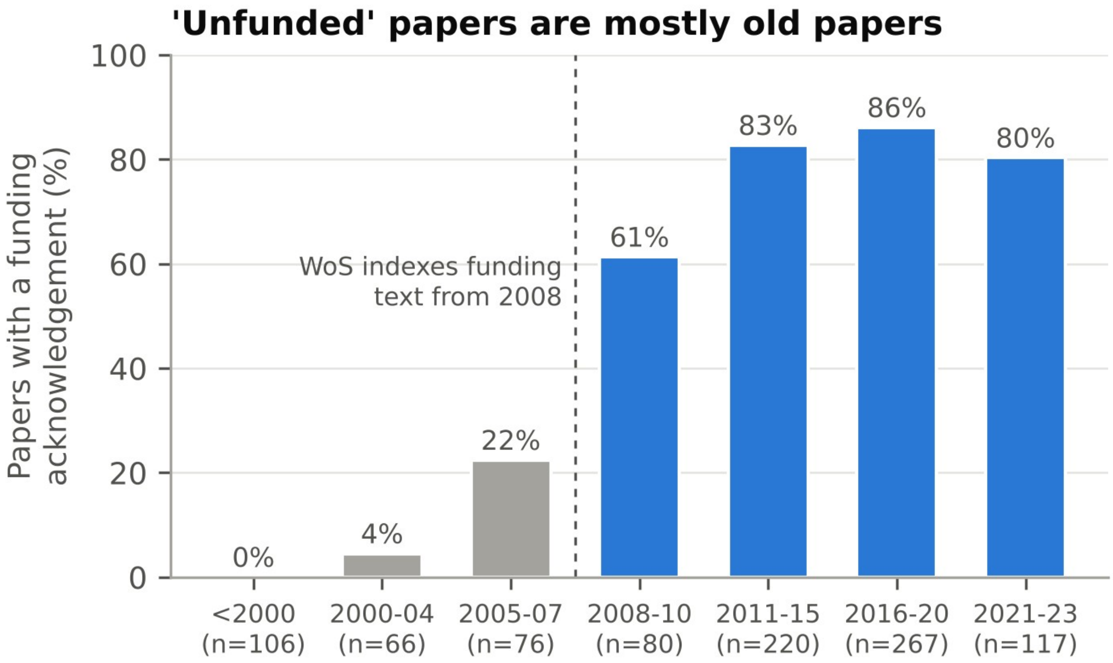

Share of all 932 records with a WoS funding record. WoS indexes funding text systematically from 2008.

##### Supplementary Figure S3. Funding and citations under different analyses

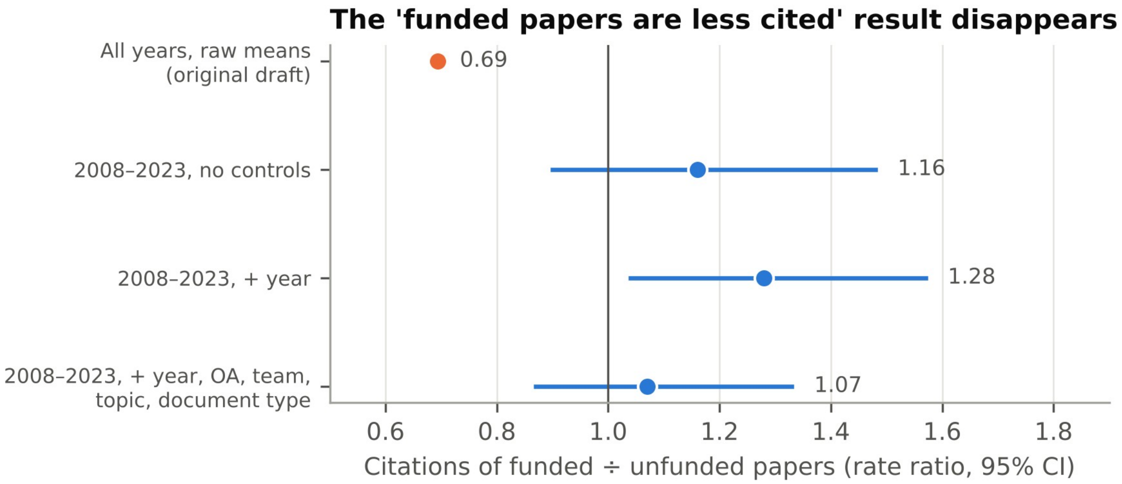

Orange: raw ratio of mean citations, all years. Blue: negative binomial rate ratios for 2008–2023, with 95% CIs.

#### Supplementary Text S1. World Bank classification

Countries were grouped by World Bank region and FY2023 income group (July 2022 classification). Venezuela was unclassified. The full table is `data/reference/country_wb_fy2023.csv`.
